# Neglected research in chronic subdural haematoma: systematic review of evidence gaps and their prioritisation by a multi-stakeholder patient and professional group

**DOI:** 10.1101/2025.07.17.25331666

**Authors:** Edward Goacher, Youssef Chedid, Munashe Veremu, William H. Cook, Matthew L. Watson, Keng Siang Lee, Orla Mantle, Vian Omar, Githmi Palahepitiya Gamage, Alexandra Lisitsyna, Matthew Kingham, Alvaro Yanez Touzet, Gideon Adegboyega, Lisa Petermann, Oliver Mowforth, Conor S. Gillespie, Daniel J. Stubbs, Benjamin M. Davies, the ICENI Clinical Practice Guidelines for Chronic Subdural Haematoma Consortia

## Abstract

**Background:** Chronic subdural haematoma (CSDH) is a common neurosurgical condition. Occurring predominantly in the elderly, its incidence is on the rise. A clinical practice guideline has been formed to offer best practice care recommendations and expose the key knowledge gaps for future research. This article presents the prioritised knowledge gaps.

**Method:** Forty-one clinical questions, across 11 themes were identified by a multidisciplinary guideline development groups. A comprehensive systematic literature search was performed on 1^st^ May 2022. 6,024 articles were screened. Themes and questions for which no relevant studies were identified are presented in this study, alongside the prioritisation of knowledge gaps a multi-disciplinary guideline group, including patients.

**Results:** In total, 19 of the 41 questions (46%) were identified as having no published literature applicable for review. The questions lay within seven themes – antithrombotic, communication, decision-making, transfer and pathway, palliative care, postop and recovery, and natural history. Within both the transfer and pathway, and the palliative care theme, 100% of the questioned posed had no applicable literature. The stakeholder group identified the top three priorities as ‘Management of antithrombotics’, ‘Benefits of Protocolised Multi-disciplinary care’, and ‘Natural history of epidemiology of non-operative CSDH’ with a cross-cutting need for a research database supported throughout.

**Conclusions:** Despite the high incidence of CSDH, almost half of the high priority research questions identified by the ICENI had studies in the published literature that could inform evidence-based recommendations. In particular, there is a paucity of literature surrounding conservatively managed cases, palliative care, and transfer pathways in CSDH.

## Introduction

Chronic subdural haematoma (CSDH) refers to a collection of ‘aged’ blood and blood breakdown products within the subdural space. It is a common form of intracranial haemorrhage, occurring predominantly in the elderly [1, 2]. In the United Kingdom (UK) alone, it is reported to occur in over 5000 people a year aged >65 [3]. Its incidence is set to rise with an ageing population and an increasing number of patients on regular antithrombotic therapy [4, 5]. It is predicted to become the most common neurosurgical operation by 2030 [6].

Surgical evacuation is recommended for patients with symptomatic CSDH [5] yet this vastly simplifies the complexity of care delivery, and the broader, perioperative challenges of treating an often frail cohort [7]. Even optimal surgical technique remains debated [8, 9] while systematic reviews in many other areas show scarce and/or low quality evidence[10]. Alternative and adjuvant treatments that have been, and continue to be, explored include dexamethasone, middle meningeal artery (MMA) embolization and tranexamic acid [10–13]. However, the role for each of these remains debated [9, 14]. Improving this evidence base led to the formation of the International Collaborative Research Initiative on Chronic Subdural Haematoma (iCORIC) study group [15]. It also underpinned the formation of The Improving Care in Elderly Neurosurgery Initiative (ICENI) in the UK, that aimed to evaluate this complex and incomplete evidence base, to produce both best practice guidelines of relevance to clinical care today, whilst also identifying priority areas for future research. [16–19].

Clinical practice guidelines are a useful tool for the identification of key knowledge gaps. The process starts by assembling multi-stakeholder working groups, with patient representation, to establish questions that require answers to inform care. [9, 16, 20] These questions then undergo systematic review and evidence appraisal. The final consensus meeting then offers an opportunity to use representative expert opinion, to prioritise these knowledge gaps.

We have previously reported on poor coverage of many non-surgical themes by systematic reviews available in the literature [10]. Here, we extend this analysis by reporting on key clinical questions in CSDH care where we identified no relevant primary literature and the prioritisation and road-mapping exercise conducted at a face-to-face consensus meeting including patients and key stakeholders.

## Methods

Guideline development methods have been reported previously [18]. The project was ethically approved [9]. For brevity, a summary of the relevant methodology for this article is included below.

### Question development

Briefly, five collaborative working groups were established – natural history and diagnosis, non-operative management, perioperative care, surgery and adjuvant care, and rehabilitation and recovery [18]. Multidisciplinary input for neurosurgery, anaesthesia, emergency care, nursing and geriatric medicine was sought for each working group [9]. Via facilitated discussion, key clinical questions that a guideline should address were identified and categorised into themes. Each question was expressed using the patient, intervention, comparison and outcome (PICO) model [21]. From the five working groups, 44 questions across 11 themes were identified in total (Supplementary Table 3).

Three questions were then removed as they related directly to the surgical technique theme and judged to have been recently and sufficiently addressed in a systematic review by Henry et al [4], leaving a total of 41 questions. Each question was deemed by the ICENI multidisciplinary group to be a ‘high priority research question’, to be answered systematically. Some of the questions identified were applicable to more than one theme (N=4).

### Literature search

A comprehensive literature search was performed on 1^st^ May 2022 of 2 databases: Medline, and Excerpta Medica Database (Embase), from inception. Reference lists and bibliographies of included articles were assessed to attempt to identify additional studies. Papers were limited to English Language. Conference abstracts, case reports, and mixed populations where it was not possible to delineate CSDH-specific outcomes (for example, a dataset containing both ASDH and CSDH data) were excluded.

Articles identified from the search were transferred to the online platform Rayyan, a repository to facilitate de-duplication and independent screening of potential records. Duplicates were removed. 11 reviewers screened 6,024 articles in total. Each article was screened by title by two blinded, independent reviewers. Articles were excluded based on a standardised initial screening criteria (Articles referring to chronic, or nonacute subdural haematoma). Included articles were then categorised dependent on their title, journal, and abstracts, according to the pre-defined ICENI themes listed in table 1. Following this, abstracts were screened, followed by full-text articles using the same process to identify manuscripts eligible for inclusion.

**Table 1.**
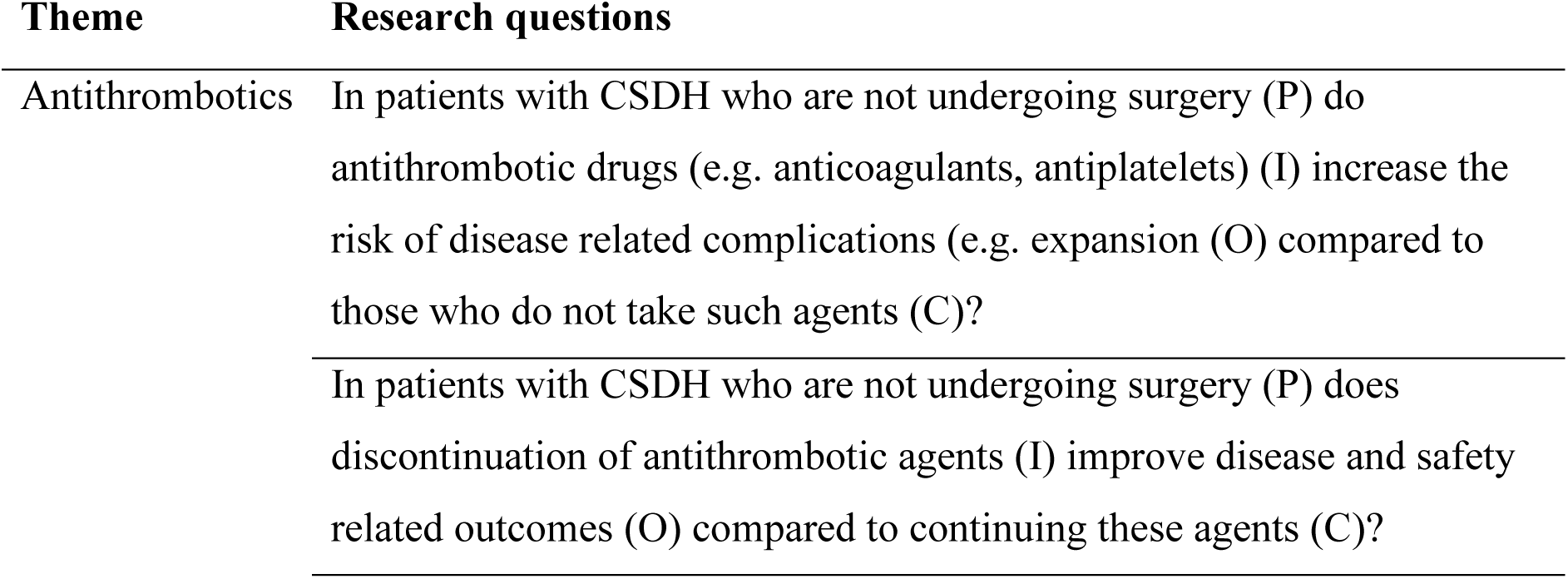
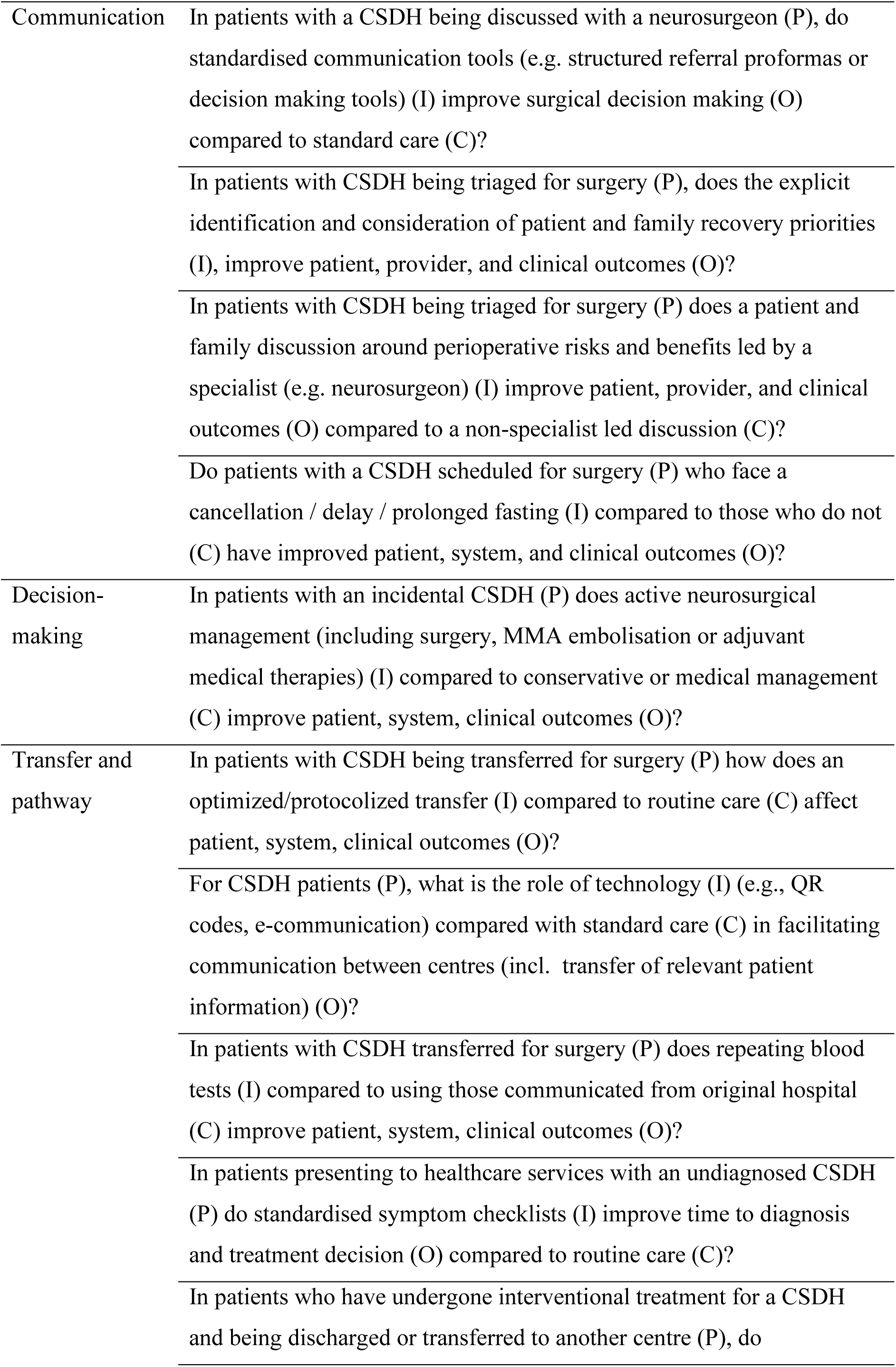
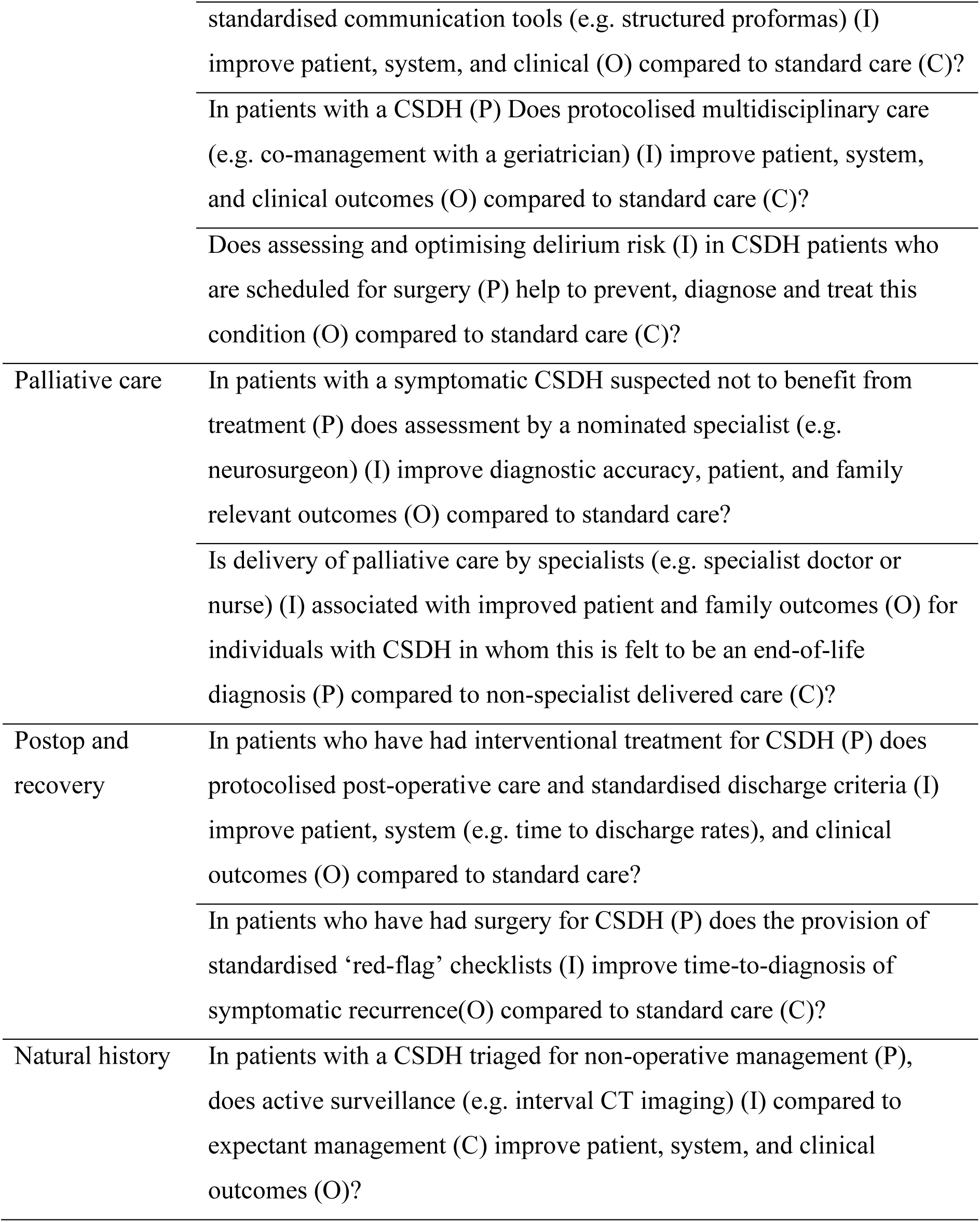
Identified questions lacking associated literature by theme. PICO model used for each question demonstrated. Abbreviations: CSDH, chronic subdural haematoma; P, population; I intervention; C comparison; O, outcome; MMA, middle meningeal artery.

Themes and questions in which no relevant studies were identified were then recorded and are presented in this study.

### Research prioritisation exercise

Draft consensus statements were proposed, by guideline development groups, where evidence was available. These where then approved, and/or iterated using a modified consensus methodology, culminating in a final face to face consensus meeting. During this consensus meeting, participants were asked to prioritise the identified knowledge gaps (aggregated into key themes), identifying both which research theme was most important to achieve, and which theme was most feasible to achieve. This could be the same theme. This exercise was conducted using a flip chart, with participants given one blue and one yellow sticky note. Participants were encouraged to add a justification to the note for their decision. The attendees’ backgrounds are described in the guideline publication [9]. Following this exercise, the top three themes (ranked by number of stick notes) were taken forward for discussion in a plenary format. The group was asked to comment on the opportunity, challenges and feasibility of answering this research priority. Alongside this, feedback on the cross-cutting theme of a research and implementation database (felt to be relevant to all the identified research themes) were also sought having distributed a proposal via a structured survey.

## Results

In total, 19 of the 41 questions (46%) were identified as having no published and applicable literature. The specific questions are displayed in Table 1. The questions lay within seven themes – antithrombotic, communication, decision-making, transfer and pathway, palliative care, postop recovery, and natural history. Seven of the questions were within the transfer and pathway theme, whilst four lay in the communication theme. Figure 1 showcases the number of published articles, stratified by research theme.

**Figure 1.**
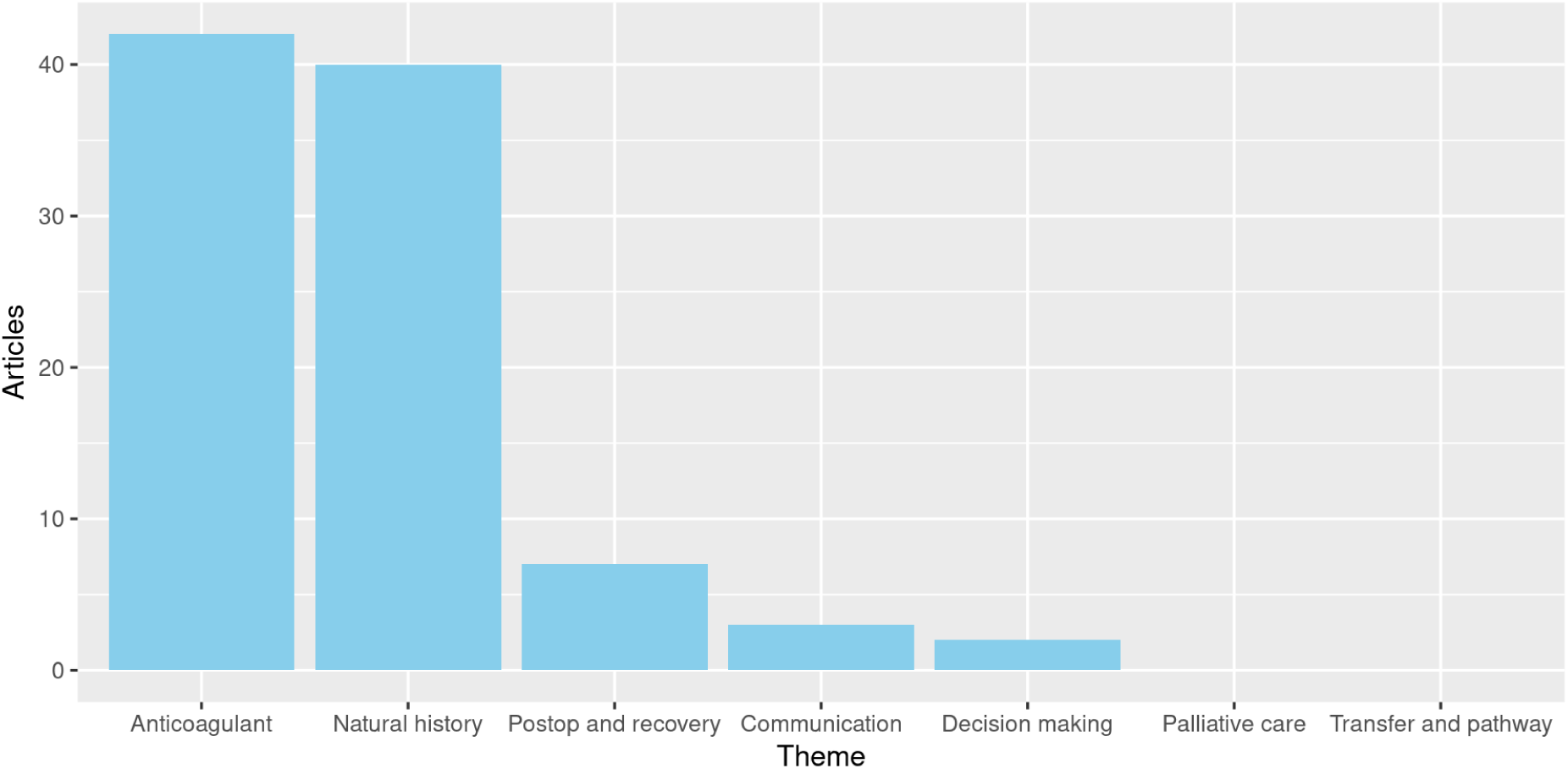
Bar chart showcasing number of published articles, stratified by each research theme.

Within both the transfer and pathway, and the palliative care theme, 100% (7/7 and 2/2, respectively) of the questions had no applicable studies. Within the communication theme, 75% (3/4) of all identified questions had no applicable studies. Fifty per cent of the identified questions within the decision-making and natural history themes had no applicable studies. Of the five questions with the postop and recovery domain, 40% (n=2) had no applicable studies. Two out of the six (33%) antithrombotic theme questions lacked usable literature. These both referred to the management and impact on antithrombotic therapy on those not undergoing surgical intervention.

The multi-stakeholder discussion results are shown in Figure 2 and Table 2. This identified the top three priorities as ‘Management of antithrombotics’, ‘Benefits of Protocolised Multi-disciplinary care’, and ‘Natural history of epidemiology of non-operative CSDH’.

**Figure 2.**
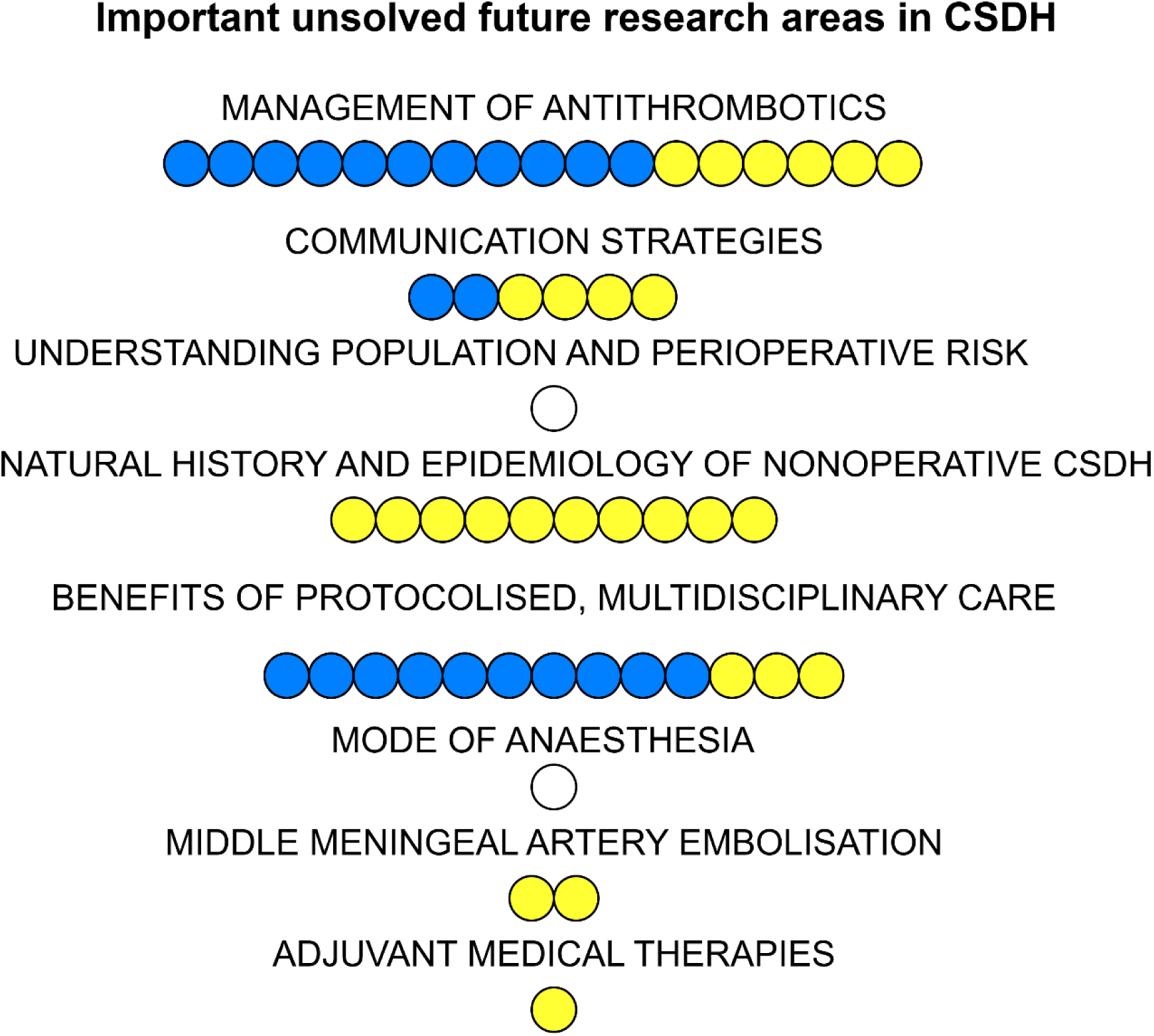
Prioritisation for unsolved research areas in CSDH, from the multi-stakeholder ICENI themes. Blue participant selected this as most important research priority. Yellow: participant selected this as most feasible research priority.

**Table 2.**
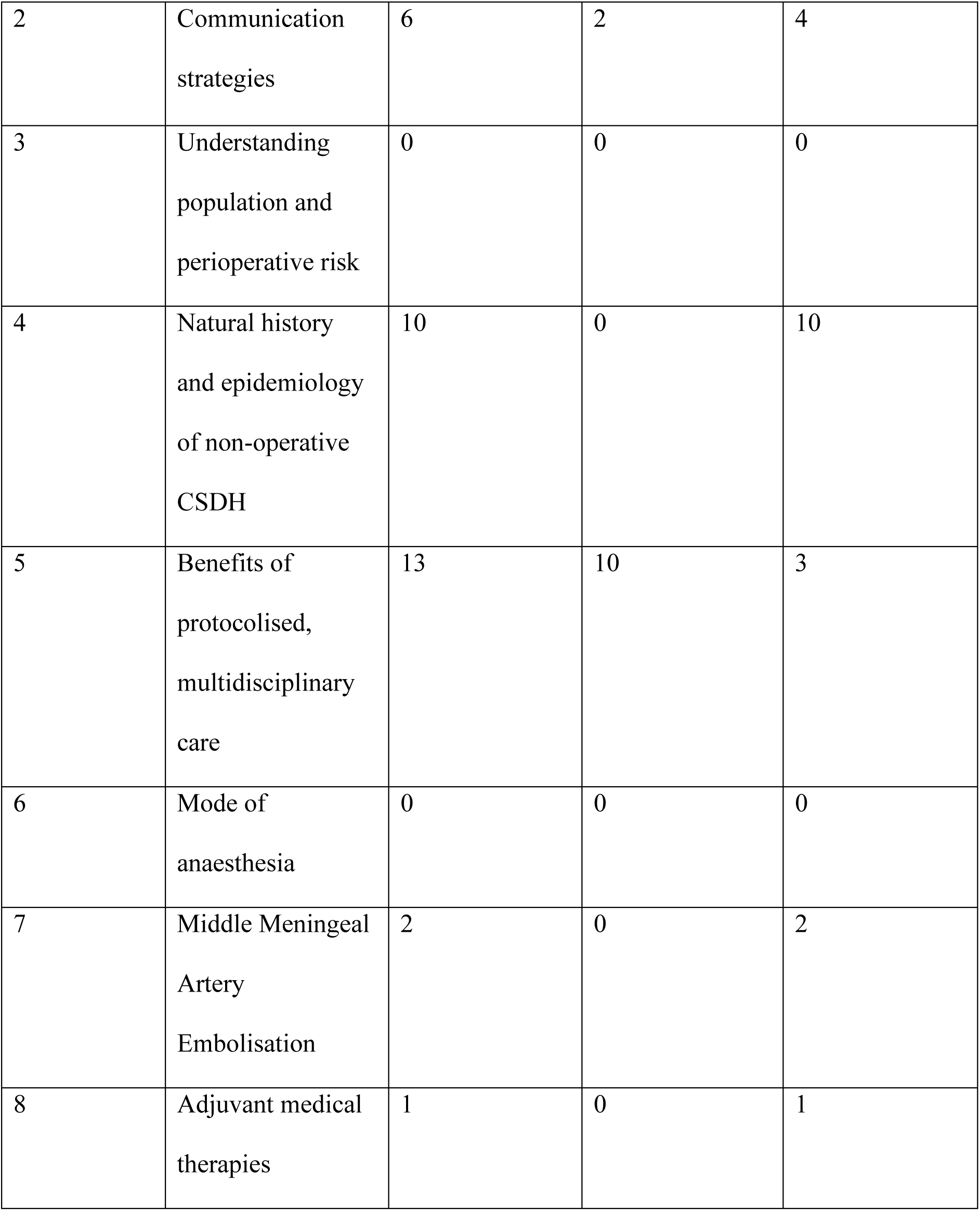
List of priorities for unsolved research areas in CSDH, from the multistakeholder consensus meeting.

| Theme number | Theme | Total number of votes | Number of 'most important priority' votes | Number of 'most feasible' votes |
| --- | --- | --- | --- | --- |
| 1 | Management of antithrombotics | 18 | 12 | 6 |
| 2 | Communication strategies | 6 | 2 | 4 |
| 3 | Understanding population and perioperative risk | 0 | 0 | 0 |
| 4 | Natural history and epidemiology of non-operative CSDH | 10 | 0 | 10 |
| 5 | Benefits of protocolised, multidisciplinary care | 13 | 10 | 3 |
| 6 | Mode of anaesthesia | 0 | 0 | 0 |
| 7 | Middle Meningeal Artery Embolisation | 2 | 0 | 2 |
| 8 | Adjuvant medical therapies | 1 | 0 | 1 |

The justifications, challenges to implementation, and potential patient impact from the discussion are shown in Table 3. For management of antithrombotics, the impact on people living with CSDH if outstanding research questions were answered was: “Reduce certainty on ongoing therapy”, “Reduce patient anxiety”, “Increase standardization of care”, “Decrease GP/hospital appointments”, “Increase safety”, “Improve patient outcomes (e.g recurrence risk/bleeding)”, and “Enhance planned theatre timing”. For natural history and epidemiology, the impact on people living with CSDH was “Avoid unnecessary operations”, “Change surgical population, “Increase alternative treatment options”, “Not impact antithrombotic”, “Address cognitive impairment”, “Impact on driving (group 2) over medication for headaches”, “Preventative measures, cosmesis”, and “General aging”. For benefits of protocolised, multidisciplinary care, the possible impact was listed as “Decrease delay”, “Improve outcomes”, “Daytime surgeries”, “Decrease variance of care”, “QALY measures/list”, and “Cross-centre/non-specialist empowerment”.

**Table 3.**
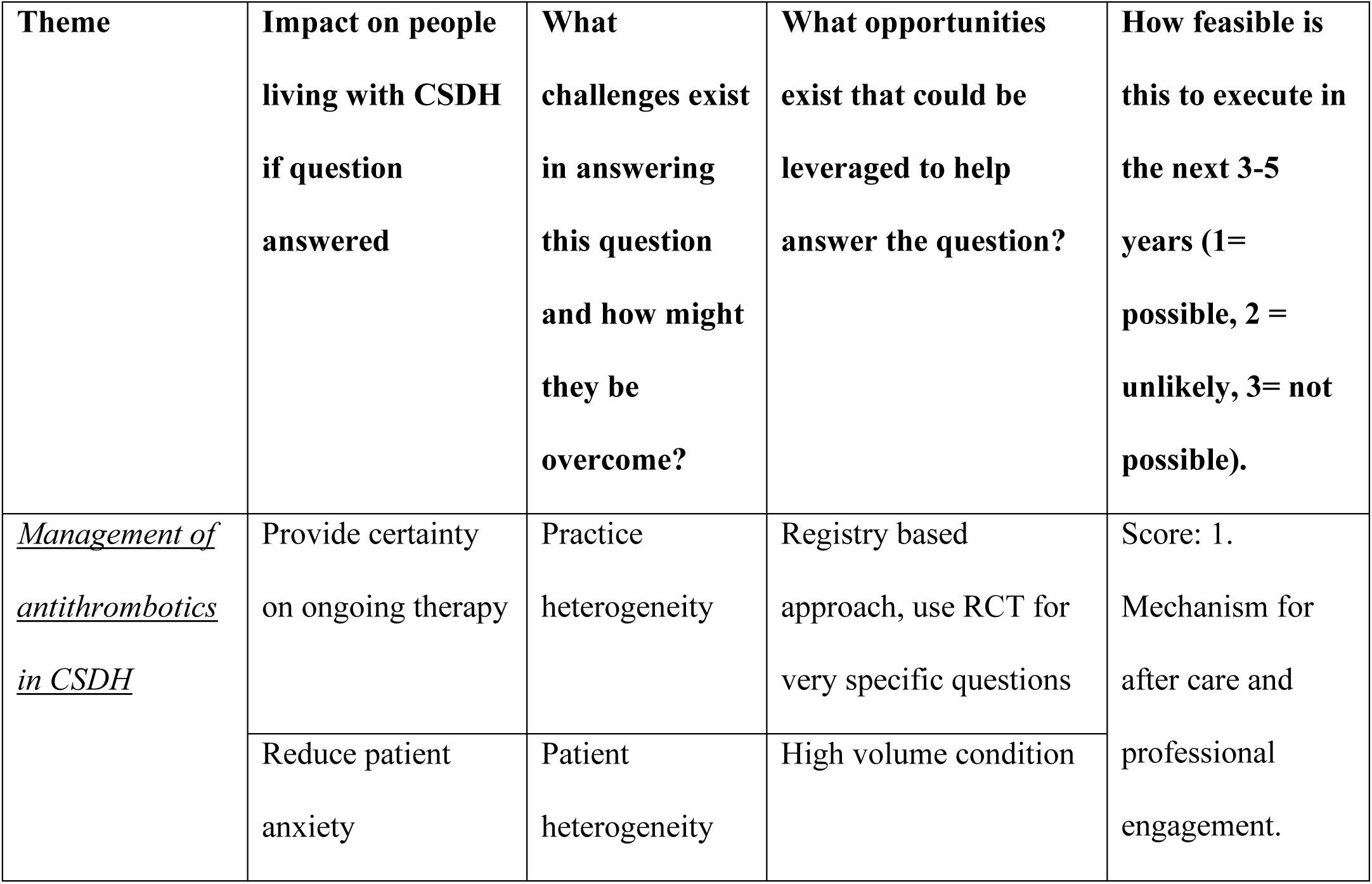
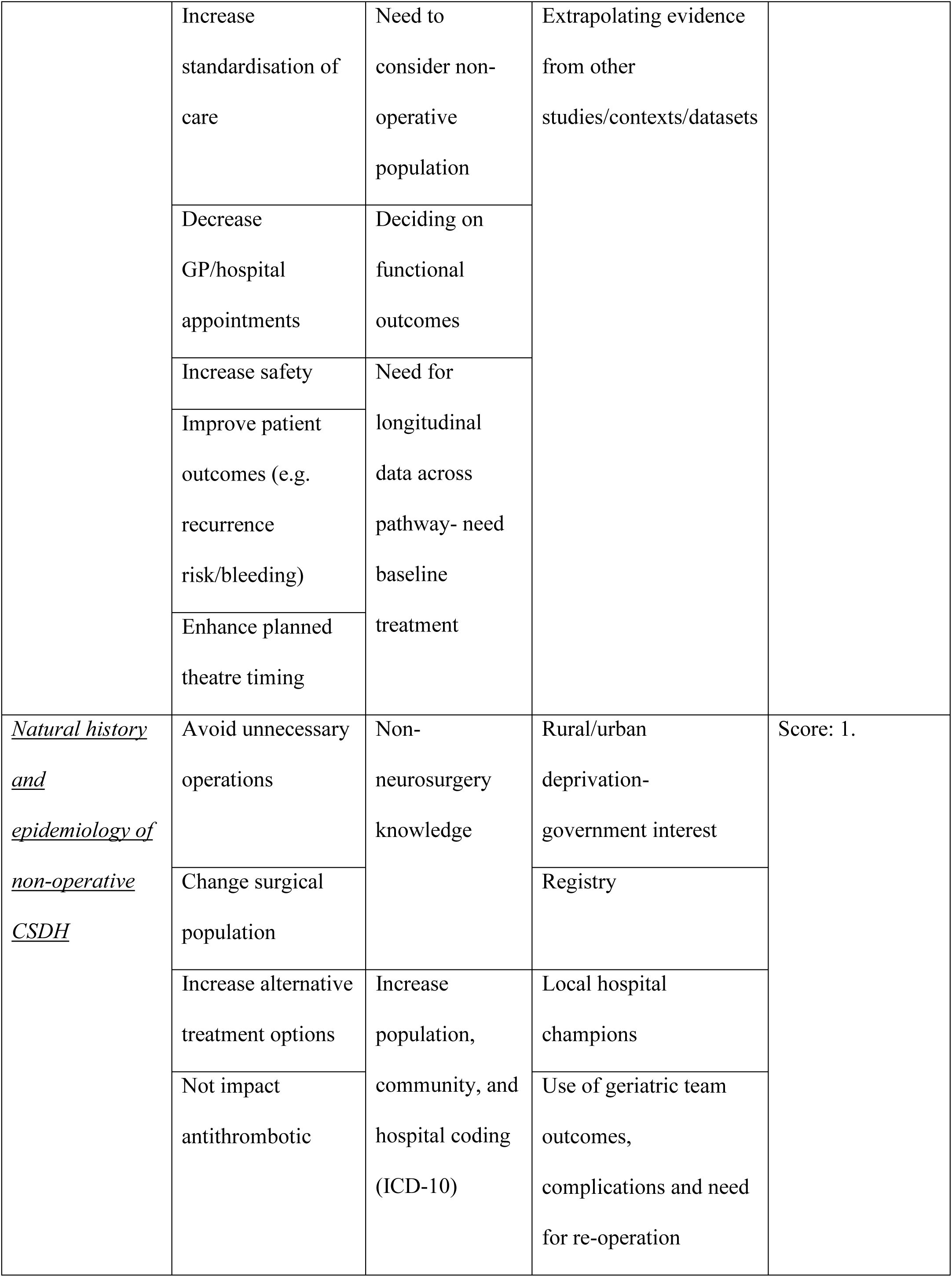
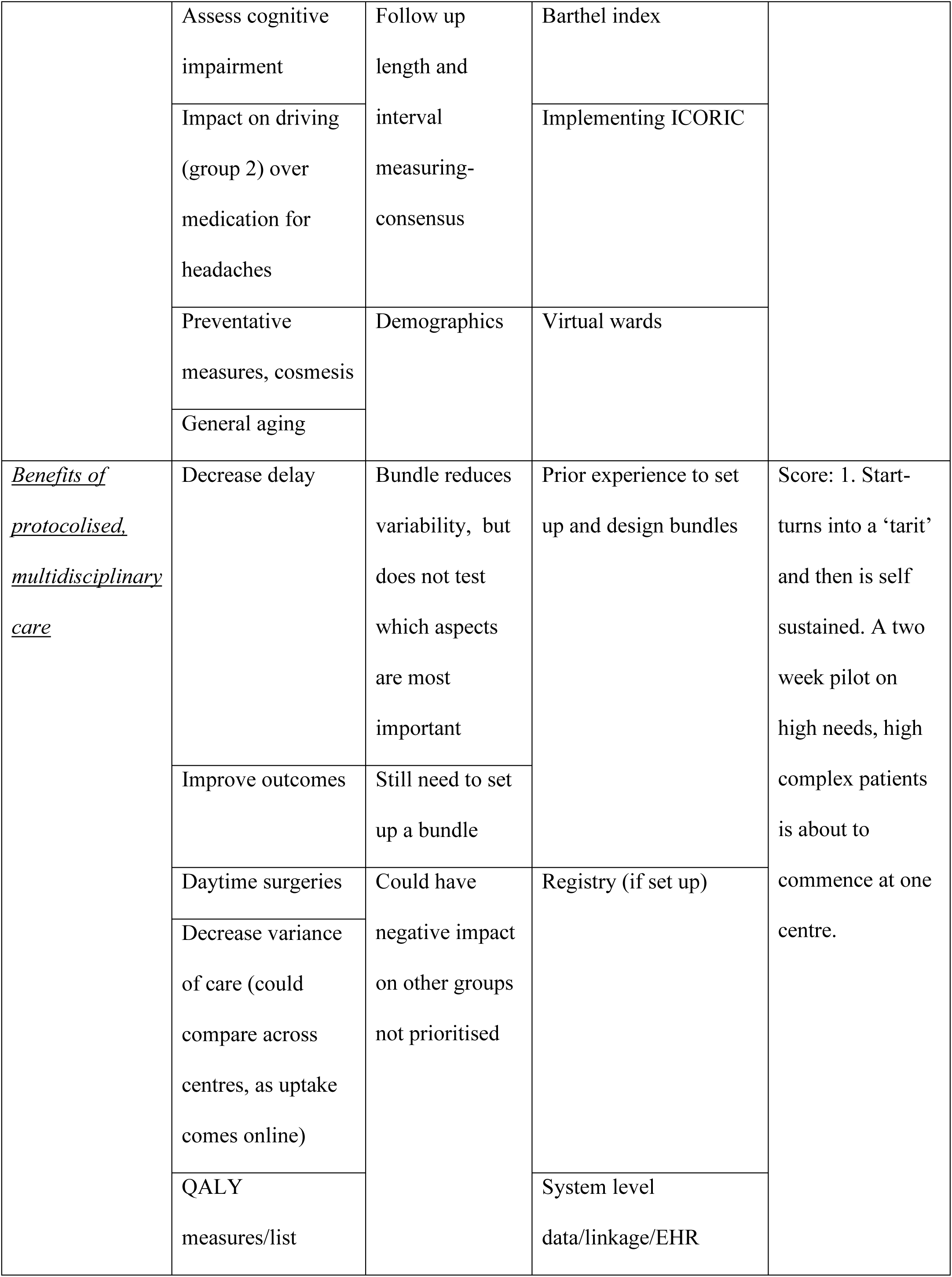
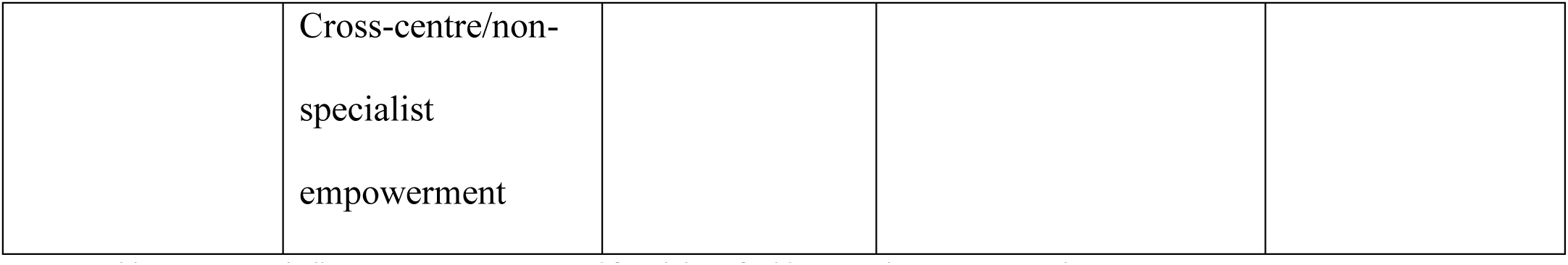
Impact, challenges, opportunities, and feasibility of addressing the top 3 research priorities in CSDH care.

### Feedback for cross-cutting theme of research database

Feedback on a CSDH research database is shown in Table 4. 96% of participants agreed a national audit or similar database for CSDH would improve patient care, and 100% agreed it would support research into CSDH care. Most of the research areas were felt to be supported by having a database. A database had support from all three patient representatives involved in the discussion.

**Table 4.** Feedback on a concept of a national CSDH database.

| Question | Participant responses |
| --- | --- |
| I think that a national audit for chronic subdural haematoma would: |  |
| Help drive improvements in patient care | 22/23 (96%) respondents agree (n = 8) or strongly agree (n = 14) |
| Help support research into the care of CsDH | 23/23 (100% respondents agree (n = 7) or strongly agree (16) |
| Be feasible to implement in practice | 14/23 (61% respondents agree (8) or strongly agree (6). The remaining participants answered 3 (n = 7) or 2 (n = 2). |
| Which of the following research areas do you think the database might help support: |  |
| Management of antithrombotics | 18/23 (78% respondents indicated this area) |
| Population and perioperative risk | 17/23 (74% respondents indicated this area) |
| Protocolized/multidisciplinary care | 14/23 (61% respondents indicated this area) |
| Role of middle meningeal artery embolisation | 16/23 (70% respondents indicated this area) |
| Communication strategies | 8/23 (35% respondents indicated this area) |
| Natural history & epidemiology | 14/23 (61% respondents indicated this area) |
| Mode of anaesthesia | 11/23 (48% respondents indicated this area) |
| Role of adjuvant medical therapies | 16/23 (70% respondents indicated this area) |
| **Patient specific questions** | **Patient responses** |
| I would be happy for anonymised information about my care to be shared with clinicians to help drive improvements in care | 3/3 (100% of respondents strongly agree) |
| I would be happy for anonymised information about my care to be shared with clinicians to help support research into CsDH | 3/3 (100% of respondents strongly agree) |
| I would be happy for information about my care to be shared with, and accessible to, clinicians who look after me in different settings if it is to help improve patient care | 3/3 (100% of respondents strongly agree) |
| I would be interested in being involved in such a project going forward: | 3/3 (100% of respondents are interested in moving forward) |

## Discussion

Despite the high incidence of CSDH, almost half of the clinical practice questions identified by the ICENI consortia as important for best practice care, had no usable studies in the current published literature. Many of these themes were highlighted by the multidisciplinary stakeholder group as important research priorities, notably the management of antithrombotic medication, understanding the natural history, and the benefits of protocolised multidisciplinary care were selected as the three most critical areas for future research. These three themes are now focused upon in this discussion.

Antithrombotics (antiplatelet and oral anticoagulant agents) are among the most commonly prescribed medication for older adults, predominantly for primary and secondary prevention of cardiovascular, stroke and thromboembolic disease [22]. Approximately 50% of patients undergoing surgery for CSDH are taking one of these medication today [23]. With an ageing and co-morbid population, their use is on the rise [5]. Given the surgical implications of antithrombotics, it is therefore not surprising that 40 studies were identified that examined the pre-, peri- and post-operative impact of antithrombotics in CSDH. That said, although pooled estimates of adverse events, such as recurrence or peri-operative veno-occlusive events could be calculated, as exclusively observational datasets, the true and comparative risk/benefits of managing this (e.g. cessation, reversal, or timing of restart) were not evaluated [5]. It therefore remains unclear whether these are truly modifiable risk factors peri-operatively.

In contrast, no studies were identified that examined the impact, nor optimal management, of antithrombotics in conservatively managed CSDH. The management of antithrombotic agents in such cases is a balancing act; the risk of further CSDH aggregation and expansion, requiring surgical treatment, versus the of veno-occlusive events posed by withholding such agents. This is further complicated by the diversity in antithrombotic agents and their indications. For example, the risk of thromboembolic events when withholding an anticoagulant in a patient with a metallic mitral heart valve is considered significantly greater than the risk in a patient on an antiplatelet agent for primary prevention of stroke [24, 25].

Each antiplatelet and anticoagulant drug exerts its antithrombotic effects by a different mechanism of action. It is already established that the risk of intracerebral haematoma is lower in direct oral anticoagulants (DOACs) than warfarin [24]. Therefore, their impact on both CSDH incidence and conservatively managed CSDH may also differ. Knowledge of the safety profile and impact of the different antithrombotic agents in conservatively managed CSDH will further inform decision making in such cases and may reduce the risk of requiring future surgical evacuation [26]. Thereby saving patients from exposure to the risks associated with surgical evacuation and preventing, potentially avoidable, costs and resource demands to health care providers [27].

With the rising incidence of CSDH, combined with the increasing prevalence and diversity of antithrombotic usage [5], their optimal management in cases of conservatively managed CSDH remains a significant unanswered question. At present, this remains a judgement call by treating clinicians and surgeons, which currently lacks any clear evidence. This was the most important research priority according to the stakeholder group. Research into this field was felt by the highest percentage of the guideline group (78%) as being aided by a national registry.

The shortage of literature relating to the communication between teams and institutions in the care of CSDH highlights the lack of, to present, a multidisciplinary team (MDT) or across institution (secondary and tertiary care) approach to its management. New guidance advocates such an approach although implementation studies are still awaited [9]. Care of the neurosurgical patient is complex and requires input from multiple specialties and team members [28]. Most patients are referred from a hospital without in-house neurosurgical services. This is particularly pertinent to CSDH. Given the complexity of the patient cohort, many have complex medical and pharmaceutical histories, including antithrombotic usage [22]. Communication between the neurosurgical, medical, anaesthetic and nursing teams is vital to optimise pre-, peri- and post-operative care of these complex patients [29].

In addition to communication amongst the MDT, the need for communication with both the patient and their families must also be noted [29]. Two clinical questions within the communication theme directly related to such communication including whether identification of patient/family recovery priorities influenced outcome and whether perioperative discussions with specialist knowledge of surgical risks and benefits improved outcomes. No related literature was found for either question. The premise for this question arose from recognition that CSDH can be an end-of-life diagnosis; the 12-month mortality after surgical treatment of a CSDH is ∼10%, increasing with advanced age. Undergoing a treatment, with a peri-operative morbidity of up to 60%, and enduring a prolonged inpatient stay may not be risks worth taking. However, it is not clear what the alternative road looks like. For example, the role of palliative care in DCM also had no supporting literature [30] [31]. Given the push in recent years towards a shared decision-making model in healthcare [32], empowering patient involvement, its impact on outcomes in CSDH remains a significant, unanswered question. Shared decision making is challenged in CSDH by the cross-institution care. Stakeholders identified this as an important area for future research.

These conversations are challenged as the natural history remains uncertain. No evidence was identified for - *In patients with an incidental CSDH, does active neurosurgical management (including surgery, MME or adjuvant medical therapies), compared to conservative or medical management, improve patient, system, clinical outcomes?* Equally no evidence was found to inform what structured non-operative management might look like. The role of surveillance imaging is unclear; CSDH can arise in people following trauma who had normal baseline CT imaging [33], and a randomised controlled trial of routine post-operative imaging was not able to better identify recurrence [34]. Understanding the natural history is therefore a very important knowledge gap for informing future care paradigms. This has been made more prescient, with the increasing introduction and availability of Middle Meningeal Artery embolization, being proposed as treatment option in pre-symptomatic CSDH [13].

Whilst the tertiary neurosurgical centre remains a focal point in treatment and triage, the route to this is varied and unstructured, as is the post-operative recovery for those referred back to local institutions. Such demands are not unique. Stroke care has been revolutionised to enable access to specialist interventional radiology service +/- surgical services. Whilst by no means perfect, these pathways have well organised triage and between institution communication and transfer policies. One of the different challenges for CSDH is that expert knowledge is not present in referring centres (stroke services are present in most secondary care settings). This is likely to place greater emphasis on communication, and perhaps telemedicine solutions. All this needs to be defined.

## Conclusion

CSDH is a common neurosurgical condition with an increasing incidence. Many questions, identified by a multi-disciplinary panel as critical to optimised patient care, remain unanswered and even unresearched. Amongst these, these experts identified three critical themes for future research: (1) Management of Antithrombotic Medication, (2) Understanding the National History and (3) Development and Implementation of Cross-Institution Multidisciplinary Care. Surgical demand for CSDH is forecast to increase by 50% over the next 20 years. These knowledge gaps need urgent progress, to ensure optimised and cost-effective healthcare.

## Data Availability

All data in the present work are contained in the manuscript

## References

1. Kolias, A.G., et al., Chronic subdural haematoma: modern management and emerging therapies. Nat Rev Neurol, 2014. 10(10): p. 570–8.

2. Miranda, L.B., et al., Chronic subdural hematoma in the elderly: not a benign disease. J Neurosurg, 2011. 114(1): p. 72–6.

3. Brennan, P.M., et al., The management and outcome for patients with chronic subdural hematoma: a prospective, multicenter, observational cohort study in the United Kingdom. J Neurosurg, 2017: p. 1–8.

4. Henry, J., et al., Management of Chronic Subdural Hematoma: A Systematic Review and Component Network Meta-analysis of 455 Studies With 103 645 Cases. Neurosurgery, 2022. 91(6): p. 842–855.

5. Brannigan, J.F.M., et al., Impact of antithrombotic agents on outcomes in patients requiring surgery for chronic subdural haematoma: a systematic review and meta-analysis. Br J Neurosurg, 2024: p. 1–8.

6. Gillespie, C.S., et al., Daytime versus out-of-hours surgery for Chronic Subdural Hematoma. J Clin Neurosci, 2024. 129: p. 110863.

7. Stubbs, D.J., B.M. Davies, and D.K. Menon, Chronic subdural haematoma: the role of peri-operative medicine in a common form of reversible brain injury. Anaesthesia, 2022. 77 Suppl 1: p. 21–33.

8. Qiu, Y., et al., Comparison of different surgical techniques for chronic subdural hematoma: a network meta-analysis. Front Neurol, 2023. 14: p. 1183428.

9. Stubbs, D.J., et al., Clinical practice guidelines for the care of patients with a chronic subdural haematoma: multidisciplinary recommendations from presentation to recovery. Br J Neurosurg, 2024: p. 1–10.

10. Gillespie, C.S., et al., How does research activity align with research need in chronic subdural haematoma: a gap analysis of systematic reviews with end-user selected knowledge gaps. Acta Neurochir (Wien), 2023. 165(7): p. 1975–1986.

11. Hutchinson, P.J., et al., A randomised, double blind, placebo-controlled trial of a two-week course of dexamethasone for adult patients with a symptomatic Chronic Subdural Haematoma (Dex-CSDH trial). Health Technol Assess, 2024. 28(12): p. 1–122.

12. Liu, J., et al., Middle Meningeal Artery Embolization for Nonacute Subdural Hematoma. N Engl J Med, 2024. 391(20): p. 1901–1912.

13. Gillespie, C.S., et al., Middle meningeal artery embolization for chronic subdural hematoma: meta-analysis of three randomized controlled trials and review of ongoing trials. Acta Neurochirurgica, 2025. 167(1): p. 166.

14. Bartek, J., et al., Multidisciplinary consensus-based statement on the current role of middle meningeal artery embolization (MMAE) in chronic SubDural hematoma (cSDH). Brain and Spine, 2024. 4: p. 104143.

15. Edlmann, E., et al., Systematic review of current randomised control trials in chronic subdural haematoma and proposal for an international collaborative approach. Acta Neurochir (Wien), 2020. 162(4): p. 763–776.

16. Stubbs, D.J., et al., Challenges and opportunities in the care of chronic subdural haematoma: perspectives from a multi-disciplinary working group on the need for change. Br J Neurosurg, 2022. 36(5): p. 600–608.

17. Adegboyega, G., et al., Seniority of Surgeon in Chronic Subdural Hematoma Recurrence: A Systematic Review and Meta-analysis. World Neurosurg, 2024. 189: p. 381–386.e1.

18. Stubbs, D.J., et al., Protocol for the development of a multidisciplinary clinical practice guideline for the care of patients with chronic subdural haematoma. Wellcome Open Res, 2023. 8: p. 390.

19. Gillespie, C.S., et al., Is information provided within chronic subdural haematoma education resources adequate? A scoping review. PLoS One, 2023. 18(4): p. e0283958.

20. Stubbs, D.J., et al., Challenges and patient outcomes in chronic subdural haematoma at the level of a regional care system A multi-centre, mixed-methods study from the East of England. Age Ageing, 2024. 53(4).

21. Schardt, C., et al., Utilization of the PICO framework to improve searching PubMed for clinical questions. BMC Med Inform Decis Mak, 2007. 7: p. 16.

22. Poon, M.T.C. and R. Al-Shahi Salman, Association between antithrombotic drug use before chronic subdural haematoma and outcome after drainage: a systematic review and meta-analysis. Neurosurg Rev, 2018. 41(2): p. 439–445.

23. Stubbs, D.J., et al., Identification of factors associated with morbidity and postoperative length of stay in surgically managed chronic subdural haematoma using electronic health records: a retrospective cohort study. BMJ Open, 2020. 10(6): p. e037385.

24. Douketis, J.D., et al., Perioperative Management of Antithrombotic Therapy: An American College of Chest Physicians Clinical Practice Guideline. Chest, 2022. 162(5): p. e207–e243.

25. Wadhera, R.K., C.E. Russell, and G. Piazza, *Cardiology patient page.* Warfarin versus novel oral anticoagulants: how to choose? Circulation, 2014. 130(22): p. e191–3.

26. Nouri, A., et al., Chronic Subdural Hematoma (cSDH): A review of the current state of the art. Brain Spine, 2021. 1: p. 100300.

27. Rauhala, M., et al., Chronic subdural hematoma-incidence, complications, and financial impact. Acta Neurochir (Wien), 2020. 162(9): p. 2033–2043.

28. Rickard, F., et al., Improving care in elderly neurosurgery initiative guideline on management of chronic subdural haematoma in older people-relevance for the geriatrician. Age Ageing, 2025. 54(1).

29. Garba, D.L., et al., Does communication between neurosurgeons and anesthesiologists improve preoperative efficiency? Clin Neurol Neurosurg, 2021. 201: p. 106461.

30. Chahine, L.M., B. Malik, and M. Davis, Palliative care needs of patients with neurologic or neurosurgical conditions. Eur J Neurol, 2008. 15(12): p. 1265–72.

31. Berthold, D., et al., Palliative care of older glioblastoma patients in neurosurgery. J Neurooncol, 2022. 157(2): p. 297–305.

32. Corell, A., et al., Shared decision-making in neurosurgery: a scoping review. Acta Neurochir (Wien), 2021. 163(9): p. 2371–2382.

33. Edlmann, E., et al., Pathophysiology of chronic subdural haematoma: inflammation, angiogenesis and implications for pharmacotherapy. J Neuroinflammation, 2017. 14(1): p. 108.

34. Schucht, P., et al., Follow-up Computed Tomography after Evacuation of Chronic Subdural Hematoma. N Engl J Med, 2019. 380(12): p. 1186–1187.

