## Supplementary material for "Neglected research in chronic subdural haematoma: systematic review of evidence gaps and their prioritisation by a multi-stakeholder patient and professional group": X

**Supplementary Table 1. Search strategies**

Original search strategy: 28^th^ April 2022. Medline via Ovid.

| Number | Search | Results |
| --- | --- | --- |
| 1 | Hematoma, Subdural, Chronic/ | 1633 |
| 2 | ((Chronic or non-traumatic or nontraumatic or spontaneous∗)) and (subdural or “sub dural”) and (haematoma∗ or hematoma∗ or haemorrhag∗ or hemorrhag∗ or bleed∗)) | 4782 |
| 3 | 1 or 2 | 4885 |

EMBASE

| Number | Search | Results |
| --- | --- | --- |
| 1 | ∗Subdural hematoma/ | 7927 |
| 2 | (chronic or non-traumatic or nontraumatic or spontaneous∗) | 2278355 |
| 3 | 1 or 2 | 3891 |
| 4 | ((“chronic subdural” or “chronic sub dural”) adj3(hematoma∗ or hematoma∗ or haemorrhag∗ or hemorrhag∗ or bleed∗)) | 3639 |
| 5 | 3 or 4 | 4635 |

**Supplementary Table 2. ICENI themes**

| Number | Theme |
| --- | --- |
| 1 | Anticoagulant |
| 2 | Decision making |
| 3 | Communication |
| 4 | Anaesthesia and Surgical scheduling |
| 5 | Transfer and pathway |
| 6 | Perioperative care |
| 7 | Palliative care |
| 8 | Postop and recovery |
| 9 | Natural history |
| 10 | Surgical technique |
| 11 | MMA Embolization |

Supplementary Table 3. Identified questions lacking associated literature by theme. PICO model used for each question demonstrated. Abbreviations: CSDH, chronic subdural haematoma; P, population; I intervention; C comparison; O, outcome; MMA, middle meningeal artery.

| **Theme** | **Research questions** |
| --- | --- |
| Antithrombotics | In patients with CSDH who are not undergoing surgery (P) do antithrombotic drugs (e.g. anticoagulants, antiplatelets) (I) increase the risk of disease related complications (e.g. expansion (O) compared to those who do not take such agents (C)? |
|  | In patients with CSDH who are not undergoing surgery (P) does discontinuation of antithrombotic agents (I) improve disease and safety related outcomes (O) compared to continuing these agents (C)? |
| Communication | In patients with a CSDH being discussed with a neurosurgeon (P), do standardised communication tools (e.g. structured referral proformas or decision making tools) (I) improve surgical decision making (O) compared to standard care (C)? |
|  | In patients with CSDH being triaged for surgery (P), does the explicit identification and consideration of patient and family recovery priorities (I), improve patient, provider, and clinical outcomes (O)? |
|  | In patients with CSDH being triaged for surgery (P) does a patient and family discussion around perioperative risks and benefits led by a specialist (e.g. neurosurgeon) (I) improve patient, provider, and clinical outcomes (O) compared to a non-specialist led discussion (C)? |
|  | Do patients with a CSDH scheduled for surgery (P) who face a cancellation / delay / prolonged fasting (I) compared to those who do not (C) have improved patient, system, and clinical outcomes (O)? |
| Decision-making | In patients with an incidental CSDH (P) does active neurosurgical management (including surgery, MMA embolisation or adjuvant medical therapies) (I) compared to conservative or medical management (C) improve patient, system, clinical outcomes (O)? |
| Transfer and pathway | In patients with CSDH being transferred for surgery (P) how does an optimized/protocolized transfer (I) compared to routine care (C) affect patient, system, clinical outcomes (O)? |
|  | For CSDH patients (P), what is the role of technology (I) (e.g., QR codes, e-communication) compared with standard care (C) in facilitating communication between centres (incl. transfer of relevant patient information) (O)? |
|  | In patients with CSDH transferred for surgery (P) does repeating blood tests (I) compared to using those communicated from original hospital (C) improve patient, system, clinical outcomes (O)? |
|  | In patients presenting to healthcare services with an undiagnosed CSDH (P) do standardised symptom checklists (I) improve time to diagnosis and treatment decision (O) compared to routine care (C)? |
|  | In patients who have undergone interventional treatment for a CSDH and being discharged or transferred to another centre (P), do standardised communication tools (e.g. structured proformas) (I) improve patient, system, and clinical (O) compared to standard care (C)? |
|  | In patients with a CSDH (P) Does protocolised multidisciplinary care (e.g. co-management with a geriatrician) (I) improve patient, system, and clinical outcomes (O) compared to standard care (C)? |
|  | Does assessing and optimising delirium risk (I) in CSDH patients who are scheduled for surgery (P) help to prevent, diagnose and treat this condition (O) compared to standard care (C)? |
| Palliative care | In patients with a symptomatic CSDH suspected not to benefit from treatment (P) does assessment by a nominated specialist (e.g. neurosurgeon) (I) improve diagnostic accuracy, patient, and family relevant outcomes (O) compared to standard care? |
|  | Is delivery of palliative care by specialists (e.g. specialist doctor or nurse) (I) associated with improved patient and family outcomes (O) for individuals with CSDH in whom this is felt to be an end-of-life diagnosis (P) compared to non-specialist delivered care (C)? |
| Postop and recovery | In patients who have had interventional treatment for CSDH (P) does protocolised post-operative care and standardised discharge criteria (I) improve patient, system (e.g. time to discharge rates), and clinical outcomes (O) compared to standard care? |
|  | In patients who have had surgery for CSDH (P) does the provision of standardised ‘red-flag’ checklists (I) improve time-to-diagnosis of symptomatic recurrence(O) compared to standard care (C)? |
| Natural history | In patients with a CSDH triaged for non-operative management (P), does active surveillance (e.g. interval CT imaging) (I) compared to expectant management (C) improve patient, system, and clinical outcomes (O)? |
